# A sociodemographic index identifies non-biological sex-related effects on insomnia in the Hispanic Community Health Study/Study of Latinos

**DOI:** 10.1101/2024.04.09.24305555

**Authors:** Natali Sorajja, Joon Chung, Carmela Alcántara, Sylvia Wassertheil-Smoller, Frank J Penedo, Alberto R Ramos, Krista M Perreira, Martha L Daviglus, Shakira F Suglia, Linda C Gallo, Peter Y Liu, Susan Redline, Carmen R Isasi, Tamar Sofer

**Affiliations:** Department of Biostatistics, Harvard T.H. Chan School of Public Health, Boston, Massachusetts, USA; Division of Sleep and Circadian Disorders, Department of Medicine, Brigham and Women’s Hospital, Boston, Massachusetts, USA; School of Social Work, Columbia University, New York, New York, USA; Department of Epidemiology & Population Health, Albert Einstein College of Medicine, Bronx, New York, USA; Department of Psychology, University of Miami, Miami, Florida, USA; Department of Neurology, University of Miami Miller wSchool of Medicine, Miami, Florida, USA; Department of Social Medicine, University of North Carolina, Chapel Hill, North Carolina, USA; Institute for Minority Health Research, University of Illinois at Chicago, Chicago, Illinois, USA; Department of Epidemiology, Rollins School of Public Health, Atlanta, Georgia, USA; Department of Psychology, San Diego State University, Chula Vista, California, USA; Division of Genetics, Lundquist Institute at Harbor-UCLA Medical Center, Torrance, CA 90502, USA; CardioVascular Institute (CVI), Beth Israel Deaconess Medical Center, Boston, Massachusetts, USA

**Keywords:** Sex differences, Gendered environment, Gendered indices, Sociodemographic sex-specific associations, Insomnia, Sleep health

## Abstract

**Background:** Sex differences are related to both biological factors and the gendered environment. To untangle sex-related effects on health and disease it is important to model sex-related differences better.

**Methods:** Data came from the baseline visit of the Hispanic Community Health Study/Study of Latinos (HCHS/SOL), a longitudinal cohort study following 16,415 individuals recruited at baseline from four study sites: Bronx NY, Miami FL, San Diego CA, and Chicago IL. We applied LASSO penalized logistic regression of male versus female sex over sociodemographic, acculturation, and psychological factors jointly. Two “gendered indices”, GISE and GIPSE, summarizing the sociodemographic environment (GISE, primary) and psychosocial and sociodemographic environment (GIPSE, secondary) associated with sex, were calculated by summing these variables, weighted by their regression coefficients. We examined the association of these indices with insomnia derived from self-reported symptoms assessed via the Women Health Initiative Insomnia Rating Scale (WHIIRS), a phenotype with strong sex differences, in sex-adjusted and sex-stratified analyses. All analyses were adjusted for age, Hispanic/Latino background, and study center.

**Results:** The distribution of GISE and GIPSE differed by sex with higher values in male individuals, even when constructing and validating them on separate, independent, subsets of HCHS/SOL individuals. In an association model with insomnia, male sex was associated with lower likelihood of insomnia (odds ratio (OR)=0.60, 95% CI (0.53, 0.67)). Including GISE in the model, the association was slightly weaker (OR=0.63, 95% CI (0.56, 0.70)), and weaker when including instead GIPSE in the association model (OR=0.78, 95% CI (0.69, 0.88)). Higher values of GISE and of GIPSE, more common in male sex, were associated with lower likelihood of insomnia, in analyses adjusted for sex (per 1 standard deviation of the index, GISE OR= 0.92, 95% CI (0.87, 0.99), GIPSE OR=0.65, 95% CI (0.61, 0.70)).

**Conclusions:** New measures such as GISE and GIPSE capture sex-related differences beyond binary sex and have the potential to better model and inform research studies of health. However, such indices do not account for gender identity and may not well capture the environment experienced by intersex and non-binary persons.

## Introduction

Sex differences are observed in many aspects of health (1–4). Modern genomics, epigenomics and other omics technologies have advanced the study of sex differences (5–7). Yet, the relationship between sex and health is more complicated than determined by binary definitions of sex as male or female. Table 1 provides an overview of terms and definitions related to sex and gender that are relevant for public health research and for this manuscript. First, sex differences are driven by a range of potential biological sex effects. Here, “biological sex effects” refer to the collection of biologically-measurable quantities related to sex, such as sex chromosome combinations, X-linked genes, reproductive organs and history, and sex hormone levels. The latter are related to, but are not completely determined by, the genetic categories of male and female sex as XY and XX sex chromosome combinations, which characterizes more than 99% of individuals (8–10). Sex hormone levels change with age, for all individuals regardless of chromosomal combinations. Thus, biological sex effects encompass a range of non-binary, time varying, effects. Second, additional health effects are due to sociocultural factors reflecting “gendered” structural and social environment, i.e., differences in occupation, income, educational attainment, family roles, etc., experienced by men, women, and non-binary individuals (11). The gendered environment of an individual is impacted by their sex assigned at birth (Figure 1), as well as by their gender identity and other gender dimensions (12). Finally, sociocultural “gender”-related factors may also intersect, affect, and interact with biological sex effects (13,14). Indeed, both biological effects and dimensions of gender have been shown to contribute to differences in cardiovascular outcomes between women and men (15). With this complexity in mind, we note that studies of sex differences in health, compared to studies examining sex- or gender-specific health determinants, still primarily focus on strata of men and women (sometimes implicitly assuming a one-to-one correspondence between binary sex and gender identity of man or woman). Thus, even when other biological and sociocultural factors are utilized, they are ultimately used to explain differences between two dichotomous groups. This manuscript addresses the issue of modeling of sex differences still within the context of two sex strata, while improving upon the dichotomization.

**Figure 1:**
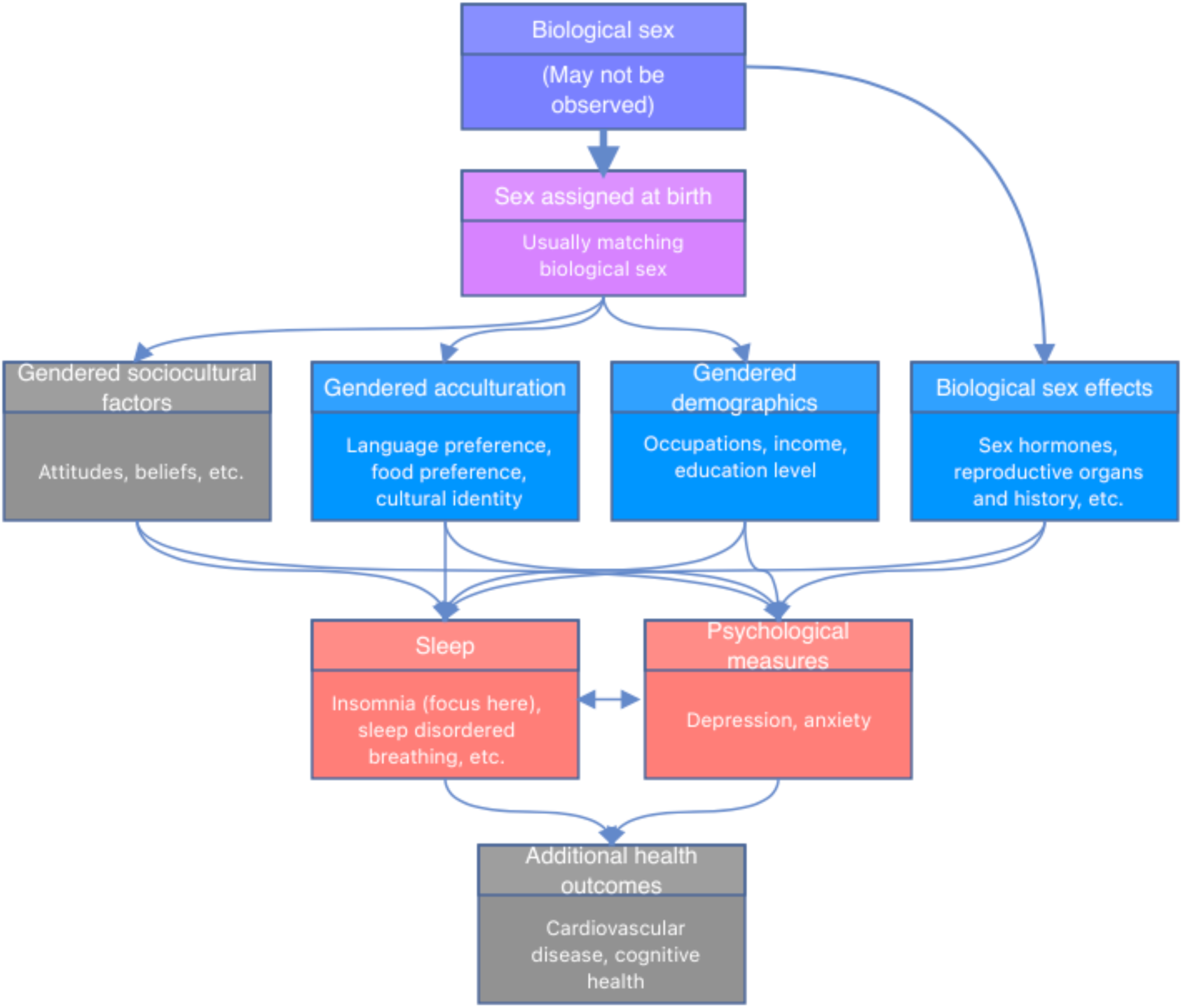
Model of sex and gender effects on sleep and health. Conceptual model of sex and gender effects on sleep. Biological sex (male, female, or other) is determined pre-birth, sex is assigned at birth, usually matching the biological sex as more than 99% of individuals are either male or female. Biological sex, where in this work we focus on binary sex, leads to biological sex-related factors, and sex assigned at birth leads to demographic, acculturation, and sociocultural (not well measured in our data, hence greyed) factors via their gendered characteristics. These may affect psychological measures and sleep, which interact. Downstream, these may affect other health outcomes (not investigated here, hence greyed).

**Table 1:** Sex- and gender-related terms used in this manuscript.

| **Term/expression** | **Meaning and context in this manuscript** |
| --- | --- |
| **Sex differences** | Differences between two groups: females and males. These groups may not be defined biologically or by gender identity, often because appropriate data are lacking, i.e. there may not be information about sex chromosomes. |
| **Biological sex effects** | Objective measures of biological factors related to sex, such as sex chromosome, sex hormones, reproductive organs and history, menopause and puberty. |
| **Sex assigned at birth** | Sex assigned at birth, typically by a physician according to observed physiological organs. |
| **Binary sex** | The categorization of individuals to male and female sex, typically with the intention to reflect underlying binary biological sex. Historically, binary sex definitions may not be accurate as individuals may be categorized to binary sex groups without an explicit check of biological definition of sex chromosomes, and are categorized instead based on gender identity or perceived gender. In research that is not focusing on sex and/or gender minorities, Individuals may be excluded from analyses focusing on binary sex. |
| **Gender dimensions** | Barr et al. (12) described four gender dimension that relates to health: identity and expression; roles and norms; relations; and power. |
| **Gender identity** | A person's sense of their own gender. In this study, we do not have data about self-reported gender identity. |
| **Gendered environment** | The idea that individuals of different gender expressions experience the world differently through differences in the environment available to them. This leads to differences in distribution of demographic and sociocultural measures between individuals of different genders (we here focus on men and women). |

A prime example of the multi-faceted sex-related effects on health is sleep, because it demonstrates strong sex differences (16,17), with some differences clearly driven by biology (18) and others likely driven by the gendered environment (19,20), including occupation, health status, and social habits (21,22). The association is also bidirectional because sleep habits/exposures such as sleep restrictions and sleep patterns related to shift work, may affect hormonal levels, including sex hormones which impact biological sex differences (23–25). Furthermore, of all behaviors, the impact of sleep is of major interest because it is intimately related to many if not all aspects of health in general (26,27). For these reasons, it is important to try to disentangle sex-related effects on sleep, and more generally, on health and disease, to better understand modifiable components of sleep health, in men, women, and all individuals.

Here, we develop gendered indices, which are data-reducing summaries of non-biological, sociodemographic measures. They are not developed to reflect nor to quantify gender identity, but rather, to summarize non-biological measures experienced by men and women independent of the direct effects of sex. We use the term “gendered” rather than, say, “sexed”, because differences in the indices distribution between male and female individuals are due to non-biological measures that are available in this dataset. These indices are constructed by using measures available in the Hispanic Community Health Study/Study of Latinos (HCHS/SOL) that are associated with binary sex. Our goal is to concisely summarize non-biological measures related to the gendered environment into a “gendered index” to be used when studying sex differences and sex-related determinants of health. We demonstrate the utility of these indices in studying sex differences in insomnia, a sleep disorder with strong sex differences – insomnia is approximately 1.5 times more common in female compared to male adults (28,29).

HCHS/SOL participants are Hispanic and Latino adults, typically first- (i.e. foreign-born with foreign-born parents) or second-generation (i.e. foreign-born with at least one U.S.-born parent) immigrants to the U.S., and often of low socioeconomic status. As such, we included measures of acculturation (i.e. of the level of assimilation, or acceptance, of the dominant U.S. culture compared to the country-of-origin culture (30)) and, in secondary analysis, psychological measures of depression and anxiety, as these are also determinants and/or correlates of both sleep health and health in general, which may show gendered patterns.

## Methods

### The Hispanic Community Health Study/Study of Latinos

Data were collected from the Hispanic Community Health Study/Study of Latinos (HCHS/SOL) epidemiologic study. The purpose of the HCHS/SOL study has been to evaluate prevalence of cardiovascular disease (CVD) risk factors, and to study the roles of and the roles CVD risk factors, socioeconomic, and genetic factors in the development of cardiovascular disease in the Hispanic/Latino population (31–33). The HCHS/SOL has data collected from 16,415 recruited participants at four different locations in the US: Miami, San Diego, Chicago, and the Bronx area of New York. Our project used data from the baseline HCHS/SOL visit which took place on 2008-2011. At baseline, sex was included in the questionnaire form and labelled “Gender” with potential responses being “Male” or “Female”. We refer to this variable as “reported sex”, because it was ascribed by the interviewer. This variable likely represents assigned sex at birth and may not describe gender identity. No options for categorization as intersex/ difference in sexual development or identification as nonbinary or transgender were provided at baseline.

However, these options have been added as part of HCHS/SOL Visit 3 which completed in early 2024 (34) and will become available in the future.

### Demographic, acculturation, and psychosocial measures

We selected sociodemographic variables that are typically associated with sex from the HCHS/SOL study. These variables included marital status, household income level, employment status, longest-held occupation type, language preference, language acculturation subscale, social acculturation subscale, both extracted from the short acculturation scale for Hispanics (35), ethnic identity score (average of two 7-point likert scale items: “I have a strong sense of belonging to my own ethnic group” and “I have a lot of pride in my ethnic group”), current health insurance status, number of years lived in the U.S., number of years of education, and psychological scores: the Spielberger Trait Anxiety Inventory scale (STAI10) measuring anxiety (36), and the Center for Epidemiological Studies Depression scale (CESD10) measuring depression symptoms (37). Because one of the CESD10 questions refers to quality of sleep (“my sleep was restless”), and we are interested in insomnia which is related, we recomputed the responses to this questionnaire, to generate “CESD9”, excluding the sleep-related question. For interpretation purposes, we re-weighted the resulting score, dividing by 9 and multiplying by 10, so it is on the same scale as the standard CESD10. In all analyses, we controlled for the baseline covariates of age, Hispanic/Latino background, and study center. We used the R *survey* package (38) to characterize the study population by sex, accounting for survey design and weights so that estimates are generalizable for the HCHS/SOL target population.

We evaluated the associations of these variables with sex using multinomial regression; each variable was treated as an outcome and individually regressed over sex as an exposure (adjusted to baseline covariates) using the *svy_VGAM* package (version 1.2) to allow for inclusion of multiple levels of a variable within the sample survey analysis framework. We next used multivariable survey logistic regression to measure the association of each sociodemographic variable with insomnia while adjusting for baseline variables, and with and without adjusting for sex. Insomnia was defined by dichotomizing the responses to the Women Health Initiative Insomnia Rating Scale (WHIIRS (39)), such that WHIIRS≥ 10 was defined as having insomnia, and no insomnia if WHIIRS<10. In these analyses insomnia was the outcome variable.

### Missing data imputation

We visualized missing patterns in data using the R *naniar* package (version 1.0.0). We performed both a complete-case analysis (primary) and sensitivity analysis based on imputed data. The sensitivity analysis was performed by imputing the data five times with the *mi* R package (version 1.1) and follow up analysis in each imputed dataset as described below. The mi function performs regression-based predictive imputation, and we used default values (other than the use of 5 imputations, whereas the default is 3).

### Development of gendered indices GISE and GIPSE

Using the sociodemographic variables in Table 2, we created two gendered indices by using least absolute shrinkage and selection operator (LASSO) regression with the *glmnet* package (version 4.1-7) to regress sex on these variables with penalization and potential variable selection. Cross-validation was performed to select the tuning parameter based on minimization of the mean-squared-error (MSE). To optimize statistical power, the entire (complete data) dataset was used to train the LASSO model. We created two indices: GISE, gendered index of sociodemographic environment (primary), which included all of the considered sociodemographic variables, and GIPSE, gendered index of psychological and sociodemographic environment (secondary), which included these variables as well as participants’ CESD9 scores and STAI10 scores. Both GISE and GIPSE were formed by extracting the fitted values from each of the two LASSO models. In secondary analysis, we also computed indices in each of the imputed datasets, and averaged the indices across imputed dataset for visualization.

**Table 2:** characteristics of the HCHS/SOL study and target population, stratified by sex.

|  |  | Female | Male |
| --- | --- | --- | --- |
| **n** |  | 9835 | 6580 |
| **Age (mean (SD))** |  | 46.54 (15.12) | 44.81 (14.84) |
| **Study site (%)** | Bronx | 2518 ( 30.3) | 1600 ( 27.6) |
|  | Chicago | 2313 ( 14.6) | 1821 ( 17.0) |
|  | Miami | 2353 ( 28.5) | 1724 ( 30.1) |
|  | San Diego | 2651 ( 26.6) | 1435 ( 25.3) |
| **Background (%)** | Dominican | 963 ( 11.5) | 510 ( 8.2) |
|  | Central American | 1049 ( 7.5) | 683 ( 7.3) |
|  | Cuban | 1250 ( 18.3) | 1098 ( 21.8) |
|  | Mexican | 4022 ( 38.1) | 2450 ( 36.6) |
|  | Puerto Rican | 1589 ( 15.3) | 1139 ( 17.1) |
|  | South American | 635 ( 5.2) | 437 ( 4.7) |
|  | More than one/Other heritage | 276 ( 4.0) | 227 ( 4.3) |
| **Education (%)** | No high school diploma or GED | 3768 ( 32.9) | 2439 ( 31.8) |
|  | At most high school diploma/GED | 2353 ( 26.3) | 1827 ( 30.2) |
|  | Greater than high school/GED | 3660 ( 40.8) | 2277 ( 38.0) |
| **Income level (%)** | Less than \$10,000 | 1587 ( 17.6) | 751 ( 11.5) |
| | \$10,001-\$20,000 | 3035 ( 33.7) | 1834 ( 29.5) |
| | \$20,001-\$40,000 | 2873 ( 32.2) | 2185 ( 34.4) |
| | \$40,001-\$75,000 | 1024 ( 12.5) | 992 ( 16.7) |
| | More than \$75,000 | 283 ( 4.0) | 363 ( 7.8) |
| **Marital status (%)** | Single | 2552 ( 31.6) | 1970 ( 37.9) |
|  | Married or living with a partner | 4726 ( 47.0) | 3710 ( 50.8) |
|  | Separated, divorced, or widow(er) | 2505 ( 21.4) | 864 ( 11.3) |
| **Employment status (%)** | Retired/not currently employed | 5264 ( 56.9) | 2689 ( 41.1) |
|  | Employed part-time(<=35 hours/week) | 1802 ( 18.8) | 926 ( 15.0) |
|  | Employed full-time(>35 hours/week) | 2596 ( 24.3) | 2832 ( 43.9) |
| **Occupation (%)** | Non-skilled worker | 2725 ( 24.0) | 2025 ( 27.5) |
|  | Service Worker | 1702 ( 18.7) | 669 ( 12.2) |
|  | Skilled Worker | 1774 ( 17.7) | 1680 ( 25.8) |
|  | Professional/technical, administrative/executive, or office staff | 1666 ( 19.6) | 699 ( 11.7) |
|  | Other occupation | 1802 ( 20.0) | 1383 ( 22.8) |
| **Current Health insurance (%)** | Yes | 5065 ( 53.2) | 3107 ( 47.7) |
| **Years in US (%)** | Less than 10 Years | 2328 ( 28.3) | 1477 ( 27.0) |
|  | 10 Years or More | 5829 ( 50.8) | 3798 ( 48.0) |
|  | US born | 1604 ( 20.9) | 1259 ( 25.1) |
| **Language preference (%)** | Spanish | 7997 ( 76.7) | 5122 ( 72.9) |
|  | English | 1838 ( 23.3) | 1458 ( 27.1) |
| **SASH** | Social acculturation subscale (mean (SD)) | 2.16 (0.60) | 2.24 (0.59) |
|  | Language acculturation subscale (mean (SD)) | 1.88 (1.12) | 2.10 (1.17) |
| **Ethnic identity score** | Mean (SD) | 3.20 (0.56) | 3.22 (0.57) |
| **Insomnia measures** | Insomnia = Yes (%) | 3424 ( 33.6) | 1578 ( 23.6) |
|  | WHIIRS (mean (SD)) | 7.68 (5.49) | 6.22 (5.03) |
| **Psychological measure** | STAI10 (mean (SD)) | 17.98 (6.10) | 16.18 (5.07) |
|  | CESD9 (mean (SD)) | 10.57 (4.97) | 9.15 (4.24) |

Proportions, means, and standard deviations (DSs) where weighted using sampling weights to represent the HCHS/SOL target population.

### Secondary analysis: verifying consistency of relationships between variables used and with reported sex

It is possible that the gendered indices are associated with sex due to overfitting to the data, as they are constructed by optimizing their association with sex. To verify that GISE and GIPSE are indeed gendered (meaning, have different association by sex), and that observed sex differences in the indices distributions are not the result of overfitting to the data, we ran a secondary analysis utilizing a training and testing set approach, by splitting the HCHS/SOL dataset into separate training and testing sets. Some of HCHS/SOL individuals were sampled from the same household. Such individuals may share similar sociodemographic and cultural environment. To ensure that the training and testing sets are independent, we verified that if an individual was in the training set, then another HCHS/SOL individual from the same household was not in the testing set. Thus, the training and testing sets were created by grouping all individuals with the same primary sampling unit (PSU) identification number into only the training or only the testing set. Finally, the training/testing sets were created using a 70%/30% split of the PSUs.

In another secondary analysis, we assessed whether the relationship among sociodemographic measures differed between males and females. We performed principal component analysis (PCA) within each sex stratum. We compared via visualization the loadings of the first male-specific and female-specific PCs to study whether the distribution of sociodemographic measures appears different between the sex strata.

### Association analysis of sex and gendered index with insomnia

We scaled the indices by dividing them by their standard deviation as computed in the complete case analysis to increase interpretability of the otherwise dimensionless variables. To measure the associations of GISE and GIPSE with insomnia, insomnia was regressed on each gendered index separately in a survey logistic regression, adjusted for the baseline covariates, including sex. The coefficient of reported sex and of the gendered indices were used to measure, correspondingly, the effects of binary sex and of the gendered environment independently of binary sex effects. The proposed interpretation of the estimated associations of GISE and GIPSE with insomnia, is that they represent sex-related effects that are explained by demographic, sociodemographic, and psychological (GIPSE) variables, on insomnia.

Because our working assumption is that the gendered indices mediate sex effects on insomnia via the sociodemographic gendered environment, we also fit a detailed association model in which insomnia was regressed on reported sex while adjusting for all the components of each of the indices, instead of GISE and GIPSE themselves. The goal was to assess whether the estimated effect of reported sex is similar in the case where only the gendered index is used compared with the case where each component variable is used individually. Finally, to analyze the relationship of insomnia and each of the indices within each sex group, we also performed sex-stratified analyses.

### Sensitivity analysis: gendered indices in imputed datasets

A new index was computed for each imputed dataset. Then, for each dataset, we repeated the association analysis with insomnia, and applied Rubin’s rule to combine the estimated effects. We also computed averaged indices across the five imputed datasets and studied their sex-specific distributions. Imputed indices may be useful for optimizing sample sizes of other analyses.

## Results

Table 2 characterizes the HCHS/SOL dataset and its target population. There were 9,835 female and 6,580 male participants. On average, female individuals in the HCHS/SOL target population were 46 years old, while male individuals were 45. In the target population, 33.6% of female individuals but only 23.6% of male individuals are estimated to have insomnia. Household income had the most missing values (n = 1,488), with social acculturation subscale (n = 729) and insomnia (n = 560) following next. There were 13,705 complete cases out of 16,415 individuals.

### Demographic, acculturation, and psychosocial measures driving sex differences in insomnia

Table 3 provides results from the multivariable regression of sociodemographic variables over sex, and the association of these variables with insomnia. These associations are further visualized in Figure 2. The results demonstrate that sociodemographic measures demonstrate sex-related patterns: all are associated with binary, reported sex. Further, most of the variables were also associated with insomnia, including when adjusting for reported sex. This supports the derivation of a combined gendered index to capture the multidimensionality of the gendered sociodemographic environment, expected to play a role in observed sex differences in insomnia.

**Figure 2:**
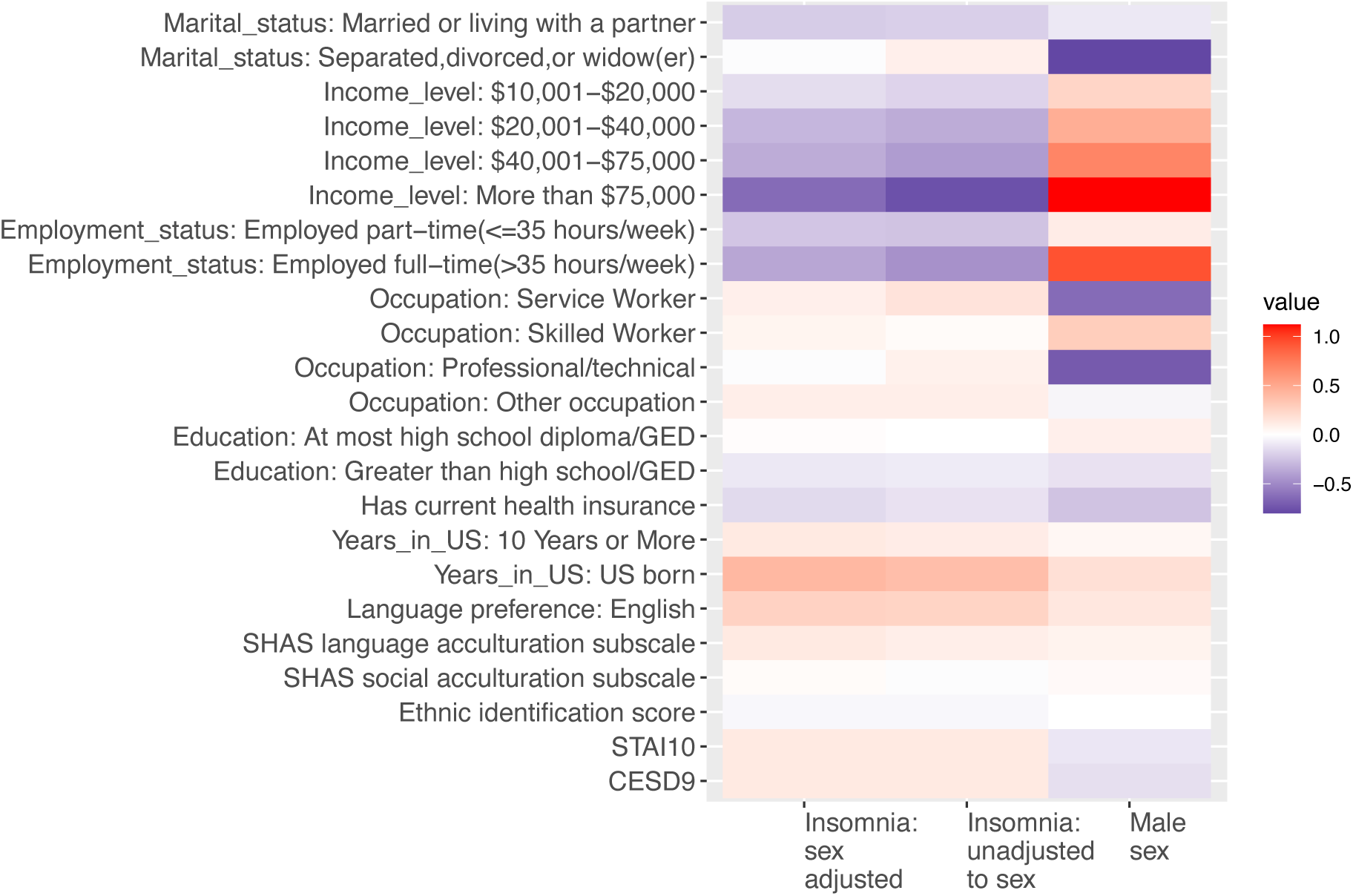
Visualization of the associations of sociodemographic and psychological variables with reported sex and insomnia. The figure visualizes the estimated associations of each of the sociodemographic and psychological variables considered with insomnia as the outcome (sex-adjusted; left column), male sex as the exposure (middle column), and insomnia as the outcome in an analysis adjusted for sex (right column). All models were adjusted for baseline covariates: age, study center, and Hispanic/Latino background. Opposite colors between sex and insomnia associations indicate that a variable is associated with higher (lower) likelihood of being male and lower (higher) likelihood of having insomnia. HCHS/SOL: Hispanic Community Health Study/Study of Latinos; SASH: Social Acculturation Scale for Hispanics; WHIIRS: Women Health Initiative Insomnia Rating Scale. STAI10: Spielberger Trait Anxiety Inventory scale based on 10 items. CESD9: Center for Epidemiological Studies Depression scale based on 9 items (excluding sleep quality-related question).

**Table 3:** Estimated association of demographic, acculturation, and psychological measures with sex and with insomnia.

|  | Beta | 95% CI | p-value | Beta | 95% CI | p-value | Beta | 95% CI | p-value |
| --- | --- | --- | --- | --- | --- | --- | --- | --- | --- |
| Characteristic | Association with male sex |  |  | Association with insomnia (sex-unadjusted) |  |  | Association with insomnia (sex-adjusted) |  |  |
| Demographic variables |  |  |  |  |  |  |  |  |  |
| Marital status (ref = Single) |  |  |  |  |  |  |  |  |  |
| Married/Living with Partner | 0.91 | (0.8,1.02) | 1.04E-01 | 0.82 | (0.72,0.94) | 3.81E-03 | 0.81 | (0.71,0.93) | 2.78E-03 |
| Divorced | 0.45 | (0.38,0.54) | 2.34E-18 | 1.09 | (0.92,1.28) | 3.26E-01 | 0.99 | (0.84,1.17) | 9.35E-01 |
| Income (ref = less than \$10000) | | | | | | | | | |
| \$10001-20000 | 1.27 | (1.08,1.5) | 3.29E-03 | 0.84 | (0.72,0.99) | 3.77E-02 | 0.87 | (0.74,1.02) | 8.7E-02 |
| \$20001-40000 | 1.59 | (1.36,1.86) | 4.81E-09 | 0.71 | (0.59,0.84) | 1.19E-04 | 0.74 | (0.62,0.89) | 1.32E-03 |
| \$40001-75000 | 1.99 | (1.67,2.38) | 1.68E-14 | 0.66 | (0.53,0.82) | 1.97E-04 | 0.71 | (0.56,0.89) | 3.08E-03 |
| More than \$75000 | 3.06 | (2.36,3.96) | 2.43E-17 | 0.47 | (0.32,0.69) | 1.33E-04 | 0.53 | (0.36,0.78) | 1.43E-03 |
| Employment (ref = Retired/unemployed) |  |  |  |  |  |  |  |  |  |
| Employed Part-Time | 1.11 | (0.96,1.28) | 1.56E-01 | 0.78 | (0.67,0.92) | 3.49E-03 | 0.79 | (0.67,0.93) | 5.35E-03 |
| Employed Full-Time | 2.54 | (2.24,2.87) | 7.44E-49 | 0.63 | (0.56,0.72) | 1.41E-12 | 0.69 | (0.61,0.79) | 1.81E-08 |
| Occupation (ref = Non-skilled worker) |  |  |  |  |  |  |  |  |  |
| Service Worker | 0.53 | (0.45,0.63) | 3E-13 | 1.17 | (0.96,1.43) | 1.1E-01 | 1.09 | (0.89,1.33) | 4.17E-01 |
| Skilled Worker | 1.33 | (1.15,1.53) | 9.29E-05 | 1.02 | (0.87,1.2) | 7.74E-01 | 1.06 | (0.9,1.24) | 4.99E-01 |
| Professional/technical | 0.49 | (0.4,0.6) | 1.15E-12 | 1.08 | (0.9,1.31) | 4.12E-01 | 0.99 | (0.82,1.2) | 9.47E-01 |
| Other Occupation | 0.96 | (0.82,1.12) | 5.94E-01 | 1.10 | (0.92,1.32) | 3.12E-01 | 1.1 | (0.91,1.31) | 3.24E-01 |
| Education (ref = No high school diploma or GED) |  |  |  |  |  |  |  |  |  |
| At most High School Diploma/GED | 1.09 | (0.95,1.24) | 2.18E-01 | 1.00 | (0.85,1.18) | 9.87E-01 | 1.01 | (0.86,1.19) | 9.1E-01 |
| Greater than high school/GED | 0.89 | (0.78,1) | 5.3E-02 | 0.92 | (0.8,1.06) | 2.51E-01 | 0.91 | (0.78,1.05) | 1.92E-01 |
| Has current health insurance coverage | 0.78 | (0.71,0.86) | 1.38E-06 | 0.89 | (0.78,1.02) | 1.02E-01 | 0.86 | (0.75,0.99) | 3.32E-02 |
| Acculturation measures |  |  |  |  |  |  |  |  |  |
| Years in the US (Ref= less than 10 years) |  |  |  |  |  |  |  |  |  |
| 10 years or more | 1.05 | (0.93,1.19) | 4.58E-01 | 1.11 | (0.97,1.28) | 1.25E-01 | 1.13 | (0.98,1.3) | 9.38E-02 |
| US born | 1.19 | (1,1.42) | 5.63E-02 | 1.45 | (1.19,1.77) | 2.09E-04 | 1.5 | (1.23,1.82) | 6.66E-05 |
| **Language preference: English** | 1.15 | (0.99,1.33) | 7E-02 | 1.28 | (1.08,1.51) | 3.8E-03 | 1.3 | (1.1,1.54) | 1.87E-03 |
| **SASH language acculturation subscale** | 0.07 | (0.05,0.09) | 3.99E-09 | 1.1 | (1.03,1.18) | 4.27E-03 | 1.13 | (1.06,1.21) | 3.62E-04 |
| **SASH social acculturation subscale** | 0.03 | (0.02,0.04) | 1.57E-05 | 0.99 | (0.88,1.11) | 8.85E-01 | 1.02 | (0.91,1.14) | 7.64E-01 |
| **Ethnic identity score** | 0.00 | (-0.01,0.01) | 7.93E-01 | 0.97 | (0.88,1.08) | 5.81E-01 | 0.97 | (0.88,1.08) | 6.1E-01 |
| **Psychological measures** |  |  |  |  |  |  |  |  |  |
| **CESD9** | -0.10 | (-0.11,-0.08) | 1.8E-30 | 1.13 | (1.12,1.14) | 5.06E-81 | 1.13 | (1.12,1.14) | 3.02E-75 |
| **STAI10** | -0.13 | (-0.15,-0.11) | 3.79E-28 | 1.13 | (1.12,1.15) | 3.57E-67 | 1.13 | (1.12,1.14) | 1.27E-62 |

The table provides the estimated associations of each considered variable with sex and insomnia from analyses adjusted for baseline covariates: age, study center, and Hispanic/Latino background. The associations were estimated separately for each variable (multiple levels of the same variables were included in the same analysis). Associations with reported sex were estimated for a multinomial regression using the svyVGAM R package, to allow for inclusion of multiple outcome levels in the same analysis. Associations with insomnia were estimated for a logistic regression with insomnia as the outcomes. Insomnia associations were further estimated in both reported sex-unadjusted and sex-adjusted analyses.

### Sociodemographic gendered indices

Table 4 provides the coefficients of the variables used in GISE and GIPSE created using penalized regression. The penalized regression model was fit using the entire data set (complete cases). Variables labeled “Not Selected” were removed by the LASSO algorithm in the variable selection process. Figure 3 visualizes the distributions of GISE and GIPSE by sex, demonstrating that male participants tend to have higher values compared to female participants.

**Figure 3:**
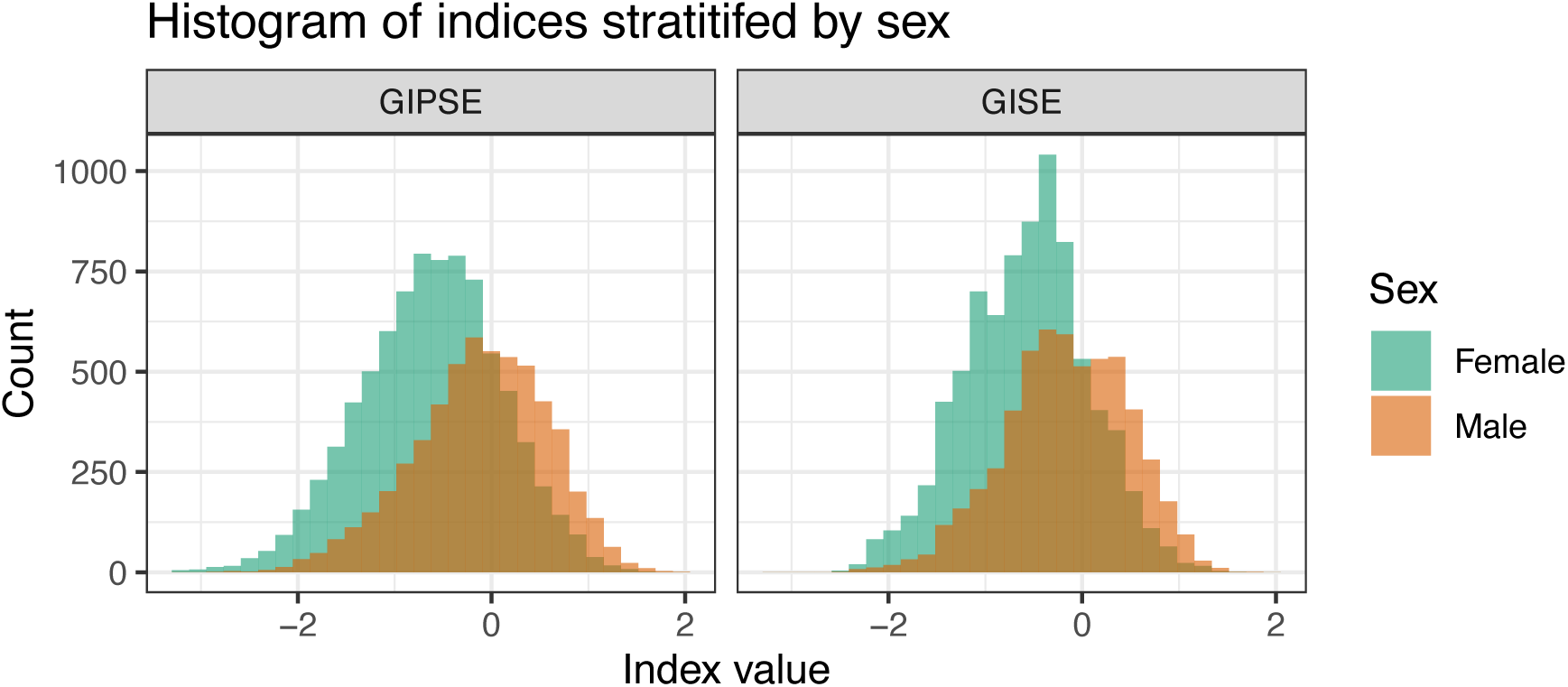
Distribution of the gendered indices in female and male participants. Distribution of the primary and secondary gendered indices by strata of reported sex. GISE: gendered index of sociodemographic environment; GIPSE: gendered index of psychological and sociodemographic environment.

**Table 4:** Gendered indices variables and coefficients.

|  | Coefficient: GISE | Coefficient: GIPSE<br>(includes<br>psychological<br>variables) |
| --- | --- | --- |
| **Demographic variables** |  |  |
| **Marital status** |  |  |
| Married or living with a partner | 0.03 | 0 |
| Separated, divorced, or widow(er) | -0.64 | -0.62 |
| **Income** |  |  |
| \$10001-20000 | 0.05 | 0 |
| \$20001-40000 | 0.13 | 0.05 |
| \$40001-75000 | 0.32 | 0.19 |
| More than \$75000 | 0.6 | 0.44 |
| **Employment** |  |  |
| Employed Part-Time | -0.09 | -0.15 |
| Employed Full-Time | 0.66 | 0.59 |
| **Occupation** |  |  |
| Service Worker | -0.62 | -0.62 |
| Skilled Worker | 0.16 | 0.17 |
| Professional/technical | -0.72 | -0.73 |
| Other Occupation | 0.06 | 0.06 |
| **Education** |  |  |
| At most High School Diploma/GED | 0.02 | 0 |
| Greater than high school/GED | -0.21 | -0.28 |
| Has Current Health insurance coverage | -0.33 | -0.32 |
| **Acculturation measures** |  |  |
| Language preference (English) | 0.3 | 0.31 |
| SASH language acculturation subscale | 0.07 | 0.06 |
| SASH social acculturation subscale | 0.15 | 0.14 |
| Ethnic identity score | 0.3 | 0.31 |
| Years live in the US (ref = Less than 10 Years) |  |  |
| Years in US: 10 Years or More | -0.08 | -0.07 |
| Years in US: US born | -0.29 | -0.28 |

| Psychological measures |  |  |
| --- | --- | --- |
| **STAI10** | NA | -0.04 |
| **CESD9** | NA | -0.03 |

Coefficients of the various variables were estimated in a LASSO penalized logistic regression of male sex over all variables jointly. Baseline variables were not used in this analysis. Thus, positive values for a binary variable suggest that it more likely to be seen in male individuals, and for a continuous variable it suggests that male individuals tend to have higher values.
GISE: gendered index of sociodemographic environment; GIPSE: gendered index of psychological and sociodemographic environment; HCHS/SOL: Hispanic Community Health Study/Study of Latinos; SASH: Social Acculturation Scale for Hispanics; WHIIRS: Women Health Initiative Insomnia Rating Scale. STAI10: Spielberger Trait Anxiety Inventory scale based on 10 items. CESD9: Center for Epidemiological Studies Depression scale based on 9 items (excluding sleep quality-related question).

In secondary analysis, we split the data to independent training and testing dataset, where we developed gendered indices on the training dataset and then constructed them in the testing dataset, and assessed whether their distribution differ by sex group. Supplementary Figure 1 visualizes the distribution of the indices in the test dataset, demonstrating that their distribution differs by reported sex, and therefore, we conclude that the gendered indices are not associated with sex only due to overfitting. Further, the figure is stratified by age groups, demonstrating that sex difference exists in the indices across adulthood. Interestingly, the median values of the gendered indices monotonically decline between the 30-40 age stratum to the 60-74 age stratum; the pattern is observed in both male and female participants.

In another sensitivity analysis, we used imputed datasets to develop gendered indices. Supplementary Figure 2 visualizes the missingness patterns in the data. Household income had the large number of observations with missing values (n = 1,488), with social acculturation subscale (n = 729) and insomnia (n = 560) following next. There were 13,705 complete cases out of the 16,415 participants. Supplementary Figure 3 visualizes the distribution of the indices based on the imputed data, averaged over the 5 imputed values for each person. In the imputed dataset, male still tend to have higher values compared to female participants, but the difference was less pronounced than in the primary analysis and in the secondary analysis of the independent test dataset within the complete-case dataset, suggesting that individuals with missing data differ from complete cases in a non-ignorable manner.

### Association of gendered indices with insomnia

Table 5 provides results from association analysis of reported sex and the developed gendered indices in multiple regression models. In a model adjusting for baseline covariates, self-reported male sex was associated with lower likelihood of having insomnia symptoms (OR=0.61, 95% CI [0.53, 0.67]). The association was attenuated after adjustment to GISE (OR=0.64, 95% CI [0.57, 0.71]), and even more so after adjustment to GIPSE (OR = 0.82, 95% CI [0.72, 0.92]). However, adjustment to the components of the gendered indices rather than to the gendered indices themselves (models 4 and 5) did not attenuate the association between male sex and insomnia to the same level, perhaps because the gendered index captures sex-related effects better.

**Table 5:** Association of reported sex and standardized gendered indices with insomnia.

| Model | Estimated male sex association |  |  | Estimated gendered index association |  |  |
| --- | --- | --- | --- | --- | --- | --- |
|  | Estimated OR | 95% CI | p-value | Estimated OR | 95% CI | p-value |
| **Model 1: adjusting for baseline covariates.** | 0.60 | (0.53,0.67) | 1.05E-16 | -- | -- | -- |
| **Model 2: adjusting for baseline covariates, GISE** | 0.63 | (0.56,0.7) | 3.9E-14 | 0.92 | (0.87,0.99) | 1.72E-02 |
| **Model 3: adjusting for baseline covariates, GIPSE** | 0.78 | (0.69,0.88) | 7.09E-05 | 0.65 | (0.61,0.7) | 2.87E-34 |
| **Model 4: adjusting for baseline covariates, components of GISE** | 0.62 | (0.55,0.7) | 3.45E-14 | -- | -- | -- |
| **Model 5: adjusting for baseline covariates, components of GIPSE** | 0.73 | (0.64,0.84) | 3.59E-06 | -- | -- | -- |
| **Analysis in male stratum** |  |  |  |  |  |  |
| **Model adjusting for baseline covariates, GISE** | -- | -- | -- | 0.93 | (0.85,1.03) | 1.58E-01 |
| **Model adjusting for baseline covariates, GIPSE** | -- | -- | -- | 0.68 | (0.62,0.75) | 9.93E-14 |
| **Analysis in female stratum** |  |  |  |  |  |  |
| **Model adjusting for baseline covariates, GISE** | -- | -- | -- | 0.92 | (0.84,0.99) | 3.42E-02 |
| **Model adjusting for baseline covariates, GIPSE** | -- | -- | -- | 0.63 | (0.58,0.69) | 8.53E-25 |

The estimated association of male sex and of the gendered indices with insomnia in various regression models. Association effects are odds ratios (OR). Thus, values lower than 1 indicate protective association. For gendered indices, ORs are per 1 standard deviation (SD) increase in the index. Male individuals tend to have higher values of the gendered indices. GISE: gendered index of sociodemographic environment; GIPSE: gendered index of psychological and sociodemographic environment.

Higher gendered indices, typical of male participants, were associated with lower insomnia likelihood (primary index OR = 0.90, 95% CI [0.68,0.85], secondary index OR = 0.62, 95% CI [0.58,0.66]) in models adjusted for sex. In separate analysis within male and female strata, GISE and GIPSE were still associated with insomnia, with similar effect size as in the combined analysis.

Supplementary Table 1 reports results from secondary analysis using individuals with imputed data as well (N=16,415 versus N=13,705 using complete cases). The association of GISE with insomnia was attenuated, and the association was not statistically significant in male individuals. The associations of GIPSE remained about the same.

## Discussion

We developed indices, GISE and GIPSE, summarizing non-biological sociodemographic, and psychological measures, such that the indices’ distributions differ by sex. Our goal was to develop a measure that will aide in distinguishing sociodemographic contributions to sex differences in health outcomes from factors that could be attributed to biological sex in Hispanic/Latino population. The gendered indices are data-driven, and they do not measure gender, as there is no specific gender-related construct that they quantify. They are “gendered” in that they have different distributions across reported male and female individuals, with typically higher values in male participants (by construction). As an exemplar, we studied the association of the gendered indices with insomnia, a phenotype that has higher prevalence in female compared to male individuals (29) with evidence relating these sex differences to both biological and sociocultural factors. Higher values of the indices were associated with lower likelihood of insomnia, even when adjusting for reported sex, and in sex-stratified analysis.

Using the GISE when modeling insomnia risk weakened somewhat the estimated association of reported sex with insomnia, suggesting that gendered sociodemographic factors explain some of the effect of reported sex on insomnia. Using GIPSE in the insomnia risk model has higher impact on the estimated effect of reported sex, further suggesting that psychological measures, depression and anxiety, have a substantial role in sex differences in insomnia.

The gendered indices are continuous rather than categorically male or female, representing a weighted aggregate of sociodemographic factors resulting in range of values. This agrees with reality, as both biological sex and gender effects are continuous. While binary sex is usually used in research, there are individuals who fall outside the standard definition based on sex chromosomes (40). Further, sex hormone levels are continuous and change throughout the reproductive cycle of women (41), throughout the life course in both men and women, and precipitously at menopause in women (42). The concept of gender is also suggested to be continuous, with measures of gender identity (43,44) and other gendered expressions and norms (45) being potentially measured by scales. Thus, binary representations of both sex and gender are limited and may not capture nuances in how sex and gender, as encompassing both biological and cultural determinants, relate to health.

Consideration of socially-constructed gendered roles and factors in relation to health and sleep is especially warranted in under-studied, diverse populations such as US-residing Hispanic/Latino individuals whose social experiences may differ in meaningful ways from populations already well-described by current literature, e.g. due to a multitude of structural and cultural components unique to this population (46–48), and whose distribution of risk for sleep disorders may not be equivalent to that of other population groups (49). For example, Kaufmann et al. (50) reported that the severity of insomnia increases with age in Hispanic adults but decreases with age in non-Hispanic White adults, strongly suggesting that assumptions regarding insomnia in Hispanic/Latino populations should not be inferred from non-Hispanic White populations. While the constructed gendered indices do not quantify any previously-specified measure of gender, we plan to use them to estimate the potential contribution of gendered sociodemographic-related burden with sleep health and other health outcomes in HCHS/SOL. As ongoing and future data collection efforts incorporate better measures of sex and gender, it will allow for more nuanced inference of the role of sex-related effects on health.

While the gendered indices replicated well to an independent subset of HCHS/SOL adults, these indices are not likely to be generalizable to other race or ethnic populations due to the cultural differences between population segments, as well as due to differences in surveys applied in different studies. The specificity of the gendered indices to HCHS/SOL (a strength) suggest that new indices will need to be tailored for other studies. Notably, we did not study potential differences in the distribution of sociodemographic measures used or in the potential resulting gendered indices between Hispanic background groups (Mexican, Cuban, Puerto Rican, etc.), which are both socioculturally diverse and exhibit different distributions of health outcomes in the HCHS/SOL (51–53). While this is certainly feasible, the indices will likely be less accurate as they will suffer from overfitting due to small sample sizes.

Strengths of this study include the use of a large cohort of diverse Hispanic/Latino adults, representing multiple Hispanic/Latino backgrounds, in the U.S., a rigorous statistical analysis that included secondary analysis validating the scores by independent training-testing data split, and imputed data analysis. There are also some limitations. The baseline HCHS/SOL dataset only included binary sex definition, and did not assess gender identity. Such data will become available in the future, based on HCHS/SOL Visit 3. Our gendered indices used household income, not the income of the individuals themselves. As household income was associated with reported sex, it is likely that either household income reporting depends on reported sex, or that household compositions in HCHS/SOL differ by reported sex. Notably, the idea of gendered indices was inspired by Smith et al. (54), who developed a gendered index in the context of labor force participation. They used a few areas of demographic measures that differ between men and women (occupation segregation, work hours, level of education, responsibility of caring for children), and included differences in income between household male and female participants in their index. By that, they interpret their index as measuring gender roles in the labor force. The indices developed here are not interpreted in such a specific context, as they have both sociodemographic (GISE) and psychological (GIPSE) components. Generally, data collection in HCHS/SOL was not specific to study sex effects, and we do not have data about additional factors that are known to have strong gendered patterns such as caregiving roles. Such measures could potentially improve the indices, but do not negate the value of the indices that have been constructed.

To summarize, we constructed two gendered indices, each summarizing sociodemographic factors, one also including psychological measures, into a single continuous variable that on average tend to have higher values in reported male individuals. The indices explain some of the association of sex with insomnia. In future work we will continue using these gendered indices to identify potentially behaviorally-modifiable aspects of sleep health and other health outcomes.

## Acknowledgements

The Hispanic Community Health Study/Study of Latinos is a collaborative study supported by contracts from the National Heart, Lung, and Blood Institute (NHLBI) to the University of North Carolina (HHSN268201300001I / N01-HC-65233), University of Miami (HHSN268201300004I / N01-HC-65234), Albert Einstein College of Medicine (HHSN268201300002I / N01-HC-65235), University of Illinois at Chicago (HHSN268201300003I / N01-HC-65236 Northwestern Univ), and San Diego State University (HHSN268201300005I / N01-HC-65237). The following Institutes/Centers/Offices have contributed to the HCHS/SOL through a transfer of funds to the NHLBI: National Institute on Minority Health and Health Disparities, National Institute on Deafness and Other Communication Disorders, National Institute of Dental and Craniofacial Research, National Institute of Diabetes and Digestive and Kidney Diseases, National Institute of Neurological Disorders and Stroke, NIH Institution-Office of Dietary Supplements. This research was partially supported by NHLBI grants R01HL161012 (to TS), R35HL135818 (to SR), and by a microgrant from the Brigham Research Institute (to TS). Additional support was provided by the Life Course Methodology Core (LCMC) at the New York Regional Center for Diabetes Translation Research (P30 DK111022).

## Data availability

HCHS/SOL data are available via a data use agreement with the HCHS/SOL Data Coordinating Center. See https://sites. cscc.unc.edu/hchs for study procedures. HCHS/SOL data are also available on the National Heart Lung and Blood Institute’s BioLINCC (Biologic Specimen and Data Repository Information Coordinating Center) repository under accession number HLB01141422a.

## Code availability

Code used in this analysis will become publicly available upon paper acceptance on the GitHub repository https://github.com/tamartsi/Gendered_indices_and_insomnia.

## Ethics statement

The HCHS/SOL was approved by the institutional review boards (IRBs) at each field center, where all participants gave written informed consent, and by the Non-Biomedical IRB at the University of North Carolina at Chapel Hill, to the HCHS/SOL Data Coordinating Center. All IRBs approving the HCHS/SOL study are: Non-Biomedical IRB at the University of North Carolina at Chapel Hill. Chapel Hill, NC; Einstein IRB at the Albert Einstein College of Medicine of Yeshiva University. Bronx, NY; IRB at Office for the Protection of Research Subjects (OPRS), University of Illinois at Chicago. Chicago, IL; Human Subject Research Office, University of Miami. Miami, FL; Institutional Review Board of San Diego State University, San Diego, CA. All methods and analyses of HCHS/ SOL participants’ materials and data were carried out in accordance with human subject research guidelines and regulations. This work was approved by the Mass General Brigham IRB and by the Beth Israel Deaconess Medical Center Committee on Clinical Investigations.

## Conflict to interests

Dr. Redline discloses consulting relationships with Eli Lilly Inc. Additionally, Dr. Redline serves as an unpaid member of the Apnimed Scientific Advisory Board, as an unpaid board member for the Alliance for Sleep Apnoea Partners, and has received loaned equipment for a multi-site study: oxygen concentrators from Philips Respironics and polysomnography equipment from Nox Medical.

## Notes

### Author Declarations

The HCHS/SOL was approved by the institutional review boards (IRBs) at each field center, where all participants gave written informed consent, and by the Non-Biomedical IRB at the University of North Carolina at Chapel Hill, to the HCHS/SOL Data Coordinating Center. All IRBs approving the HCHS/SOL study are: Non-Biomedical IRB at the University of North Carolina at Chapel Hill, Chapel Hill, NC: Einstein IRB at the Albert Einstein College of Medicine of Yeshiva University, Bronx, NY; IRB at Office for the Protection of Research Subjects (OPRS), University of Illinois at Chicago. Chicago, IL; Human Subject Research Office, University of Miami. Miami, FL; Institutional Review Board of San Diego State University, San Diego, CA. All methods and analyses of HCHS/ SOL participants' materials and data were carried out in accordance with human subject research guidelines and regulations. This work was approved by the Mass General Brigham IRB and by the Beth Israel Deaconess Medical Center Committee on Clinical Investigations.

