## Supplementary materials for "A sociodemographic index identifies non-biological sex-related effects on insomnia in the Hispanic Community Health Study/Study of Latinos"

### Supplementary Figures

#### Supplementary Figure 1: Distribution of gendered indices in independent test dataset

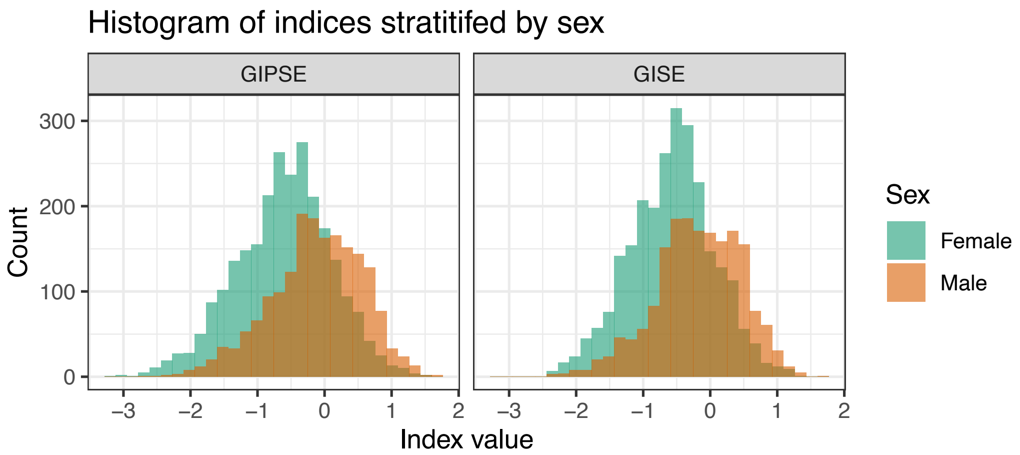

Histograms of the GISE and GIPSE, stratified by age group, in a dataset composed of 4,070 individuals who are all from different primary sampling units of the 9,596 individuals who were used to train the indices.

GISE: gendered index of sociodemographic environment; GIPSE: gendered index of psychological and sociodemographic environment.

#### Supplementary Figure 2: Missingness patterns

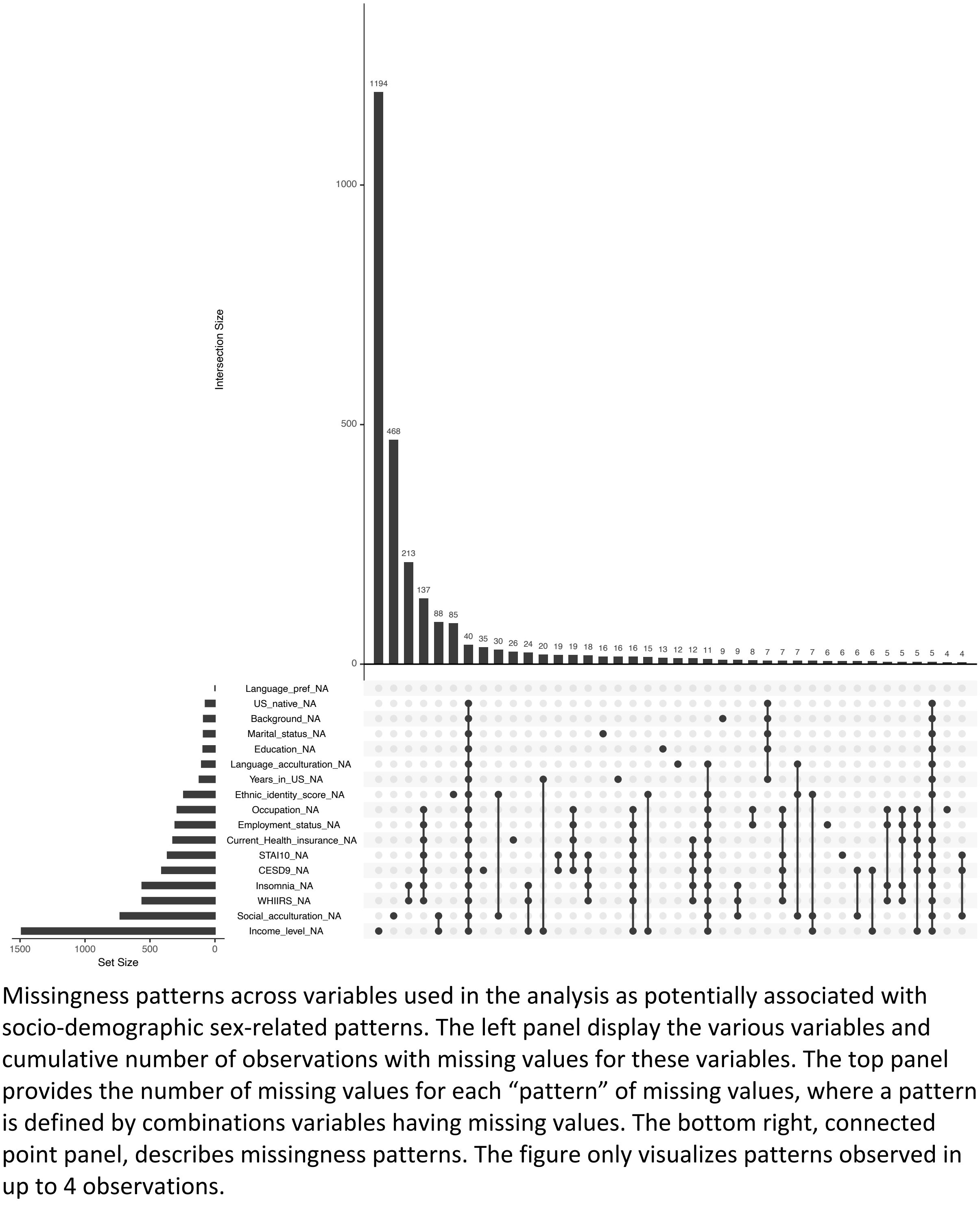

Missingness patterns across variables used in the analysis as potentially associated with socio-demographic sex-related patterns. The left panel display the various variables and cumulative number of observations with missing values for these variables. The top panel provides the number of missing values for each “pattern” of missing values, where a pattern is defined by combinations variables having missing values. The bottom right, connected point panel, describes missingness patterns. The figure only visualizes patterns observed in up to 4 observations.

#### Supplementary Figure 3: Distribution of gendered indices in imputed data

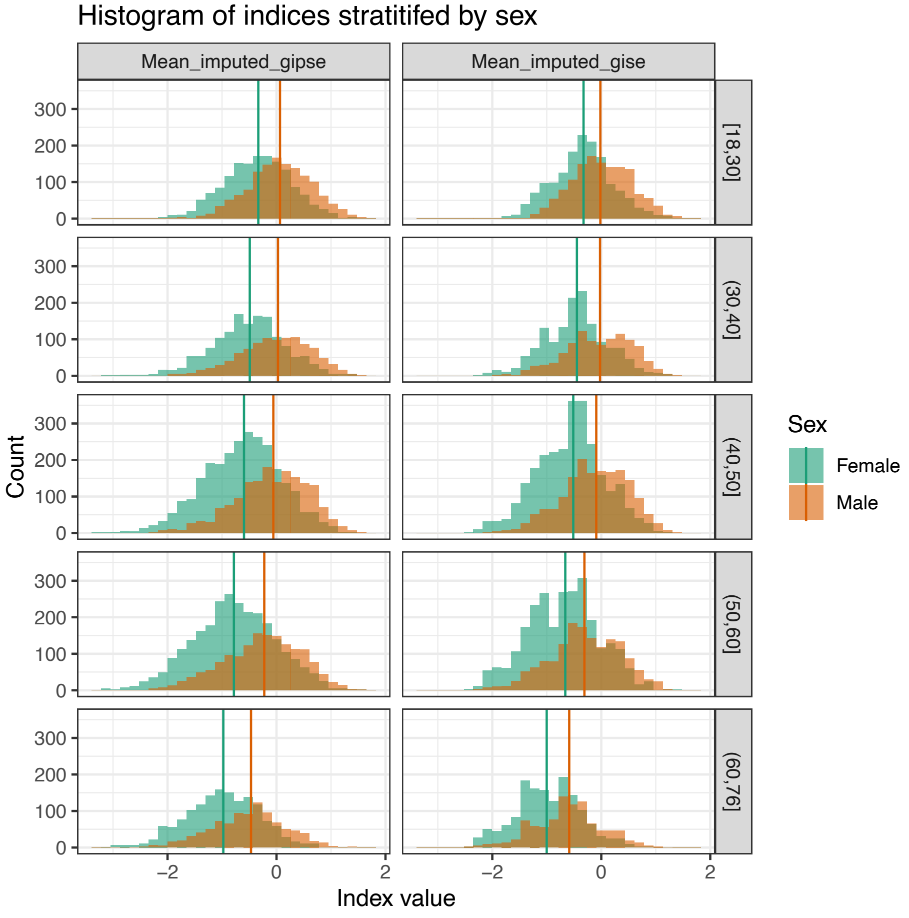

Histograms of GISE and GIPSE computed based on the imputed dataset. The lines correspond to median values of the indices within male and female participants in the relevant age group.

GISE: gendered index of sociodemographic environment; GIPSE: gendered index of psychological and sociodemographic environment.

#### Supplementary Figure 4: Principal component analysis of demographic, acculturation, and psychological variables demonstrate similar relationship between variables in males and females

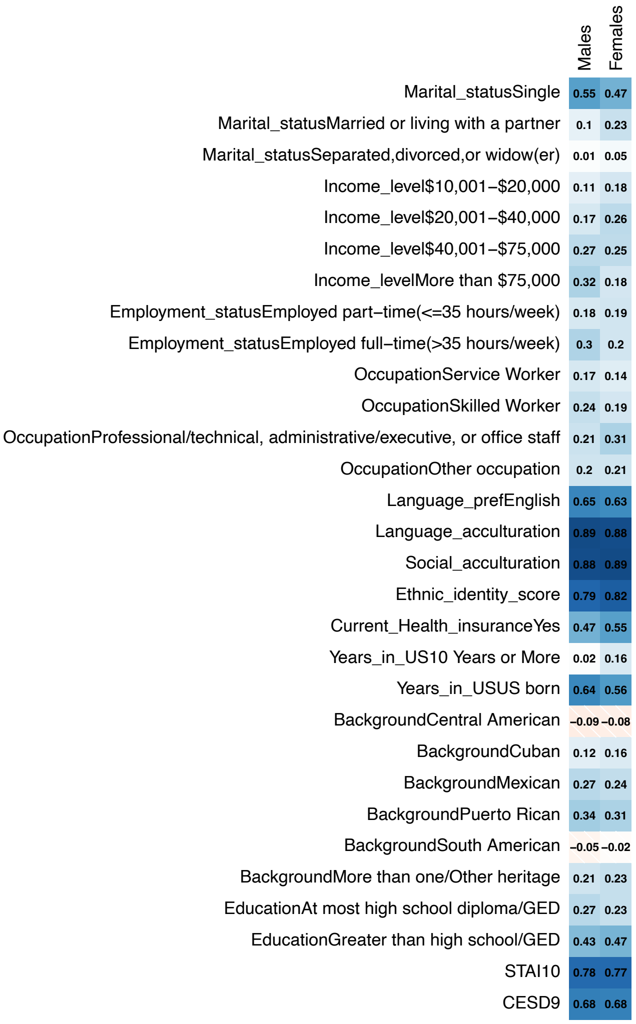

Loadings of the first principal components of sociodemographic variables used, computed separately in males and in females only.

### Supplementary Tables

#### Supplementary Table 1: Association analysis of sex and gendered index with insomnia using imputed data

| Model | Estimated male sex effect | | | Estimated gendered index effect | | |
|---|---|---|---|---|---|---|
| | Estimated effect | 95% CI | p-value | Estimated effects | 95% CI | p-value |
| **Model 1: adjusting for baseline covariates.** | 0.61 | (0.55,0.67) | 2.66E-20 | NA | NA | NA |
| **Model 2: adjusting for baseline covariates, GISE** | 0.63 | (0.57,0.70) | 6.57E-17 | 0.93 | (0.88,0.99) | 1.96E-02 |
| **Model 3: adjusting for baseline covariates, GIPSE** | 0.79 | (0.71,0.88) | 3.33E-05 | 0.65 | (0.61,0.69) | 1.26E-45 |
| **Model 4: adjusting for baseline covariates, components of GISE** | 0.63 | (0.56,0.70) | 5.24E-17 | NA | NA | NA |
| **Model 5: adjusting for baseline covariates, components of GIPSE** | 0.75 | (0.66,0.84) | 1.38E-06 | NA | NA | NA |
| **Analysis in male stratum** | | | | | | |
| **Model adjusting for baseline covariates, GISE** | NA | NA | NA | 0.95 | (0.87,1.03) | 2.24E-01 |
| **Model adjusting for baseline covariates, GIPSE** | NA | NA | NA | 0.68 | (0.62,0.75) | 2.9E-16 |
| **Analysis in Female stratum** | | | | | | |
| **Model adjusting for baseline covariates, GISE** | NA | NA | NA | 0.92 | (0.85,0.99) | 3.12E-02 |
| **Model adjusting for baseline covariates, GIPSE** | NA | NA | NA | 0.63 | (0.58,0.68) | 5.42E-33 |
